# Evaluating COVID-19 reporting data in the context of testing strategies across 31 LMICs

**DOI:** 10.1101/2021.02.11.21251590

**Authors:** Mollie M. Van Gordon, Kevin A. McCarthy, Joshua L. Proctor, Brittany L. Hagedorn

## Abstract

**Background:** COVID-19 case counts are the predominant measure used to track epidemiological dynamics and inform policy decision-making. Case counts, however, are influenced by testing rates and strategies, which have varied over time and space. A method to consistently interpret COVID-19 case counts in the context of other surveillance data is needed, especially for data-limited settings in low- and middle-income countries (LMICs).

**Methods:** We leverage statistical analyses to detect changes in COVID-19 surveillance data. We apply the pruned exact linear time change detection method for COVID-19 case counts, number of tests, and test positivity rate over time. With this information, we categorize change points as likely driven by epidemiological dynamics or non-epidemiological influences such as noise.

**Findings:** Higher rates of epidemiological change detection are more associated with open testing policies than with higher testing rates. We quantify alignment of non-pharmaceutical interventions with epidemiological changes. LMICs have the testing capacity to measure prevalence with precision if they use randomized testing. Rwanda stands out as a country with an efficient COVID-19 surveillance system. Sub-national data reveal heterogeneity in epidemiological dynamics and surveillance.

**Interpretation:** Relying solely on case counts to interpret pandemic dynamics has important limitations. Normalizing counts by testing rate mitigates some of these limitations, and open testing policy is key to efficient surveillance. Our findings can be leveraged by public health officials to strengthen COVID-19 surveillance and support programmatic decision-making.

**Funding:** This publication is based on models and data analysis performed by the Institute for Disease Modeling at the Bill & Melinda Gates Foundation.

**Research in Context:** *Evidence before this study:* We searched for articles on the current practices, challenges, and proposals for COVID-19 surveillance in LMICs. We used Google Scholar with search terms including “COVID surveillance.” Existing studies were found to be qualitative, anecdotal, or highly location-specific.

*Added value of this study:* We developed a quantitative method that makes use of limited information available from LMICs. Our approach improves interpretation of epidemiological data and enables evaluation of COVID-19 surveillance dynamics across countries.

*Implications of all the available evidence:* Our results demonstrate the importance of open testing for strong surveillance systems, bolstering existing anecdotal evidence. We show strong alignment across LMICs between non-pharmaceutical interventions and epidemiological changes. We demonstrate the importance of considering sub-national heterogeneity of epidemiological dynamics and surveillance.

## 1. Introduction

The virus known as SARS-CoV-2 was first identified in Wuhan, China in December 2019. Since then, countries have scrambled to monitor the severity and trajectory of the COVID-19 outbreak and to control its progression using non-pharmaceutical interventions (NPIs). Disease surveillance has mostly relied on case counts to inform public health policies (WHO 2020). There has not, however, been a robust evaluation of case counts as a metric for epidemiological dynamics, nor the varied surveillance approaches used to track disease trajectories.

Case-based surveillance systems have known weaknesses, including the strong influence of testing rates, which vary widely across space and time (Haider et al. 2020). Case counts can be inconsistently measured, testing capacity limited, and eligibility policies variable. It is critical to understand the limitations of available data and to identify metrics that are robust to these challenges, particularly for low- and middle-income countries (LMICs).

There is general recognition that surveillance system performance can be a challenge in LMICs, and that understanding disease surveillance is key to system improvement and production of representative data (Petti et al. 2006). Existing efforts to evaluate LMIC surveillance systems, however, are largely qualitative, country-specific, or based on commentary (Ibrahim 2020; Farahbakhsh et al. 2020; Alwan 2020). Further, most national-level studies of NPI impacts focus on high-income countries, but there is evidence that these insights cannot be readily generalized to LMIC settings (Brauner et al. 2020; Flaxman et al. 2020; Hsiang et al. 2020; Dehning et al. 2020; Chen et al. 2020; Islam et al. 2020; Haider et al. 2020). This leaves an important knowledge gap in understanding how to evaluate and interpret COVID-19 epidemiological data from LMICs.

To address the gap in systematic interpretation and evaluation methods, we leverage statistical analysis techniques to detect changes in underlying properties of COVID-19 time series surveillance data across 31 LMICs. With this information, we categorize detected change points as likely driven by epidemiological changes or non-epidemiological influences such as noise. This provides a quantitative and automated approach to analyzing epidemiological surveillance data. We make use of imperfect information despite data weaknesses, deriving insights from information available in LMICs that may otherwise be overlooked. The approach is fast and highly portable, well suited to looking across countries, and has minimal data requirements.

In this article, we first present the methods for our analysis, including the statistical model, change point categorization, and evaluation of epidemiological change co-occurrence with NPIs. We follow with validation of our method, the usefulness of open testing, comparisons of country surveillance characteristics, and consideration of sub-national dynamics. Finally, we elaborate on the significance of our results, broader conclusions, and relevance for public health applications.

## 2. Methods

The methods are outlined in Figure 1 for two example countries: South Africa and Bangladesh. Details about each step are presented in the following subsections.

**Figure 1:**
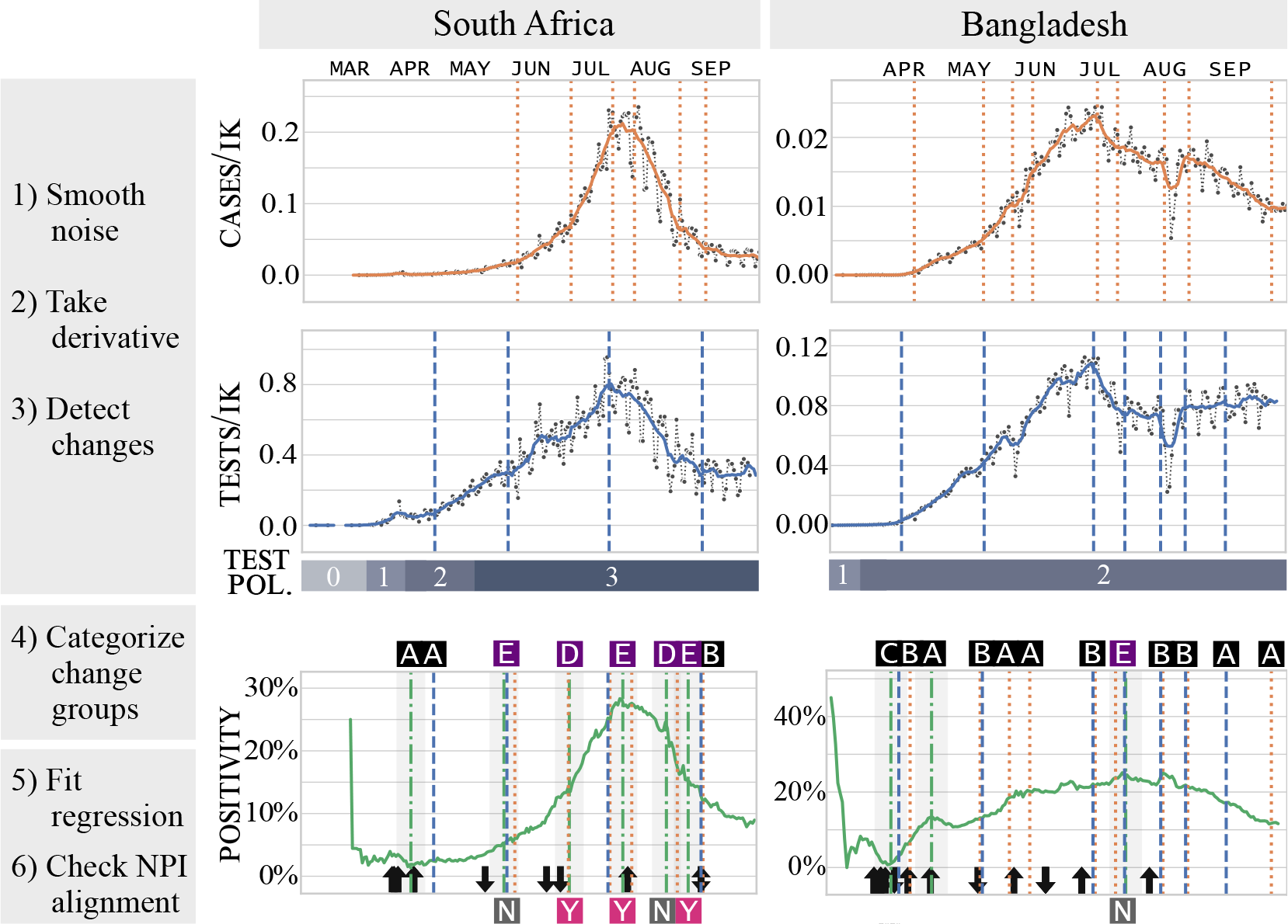
Methods Overview. Time series for cases (orange), tests (blue) and positivity (green) for case study countries South Africa and Bangladesh. Cases and tests are plotted in units per thousand people. Vertical lines indicate detected change points on each series. National changes in testing policy are shown as blue bars; see Section 2.1 for policy descriptions. Positivity change points are overlaid with case and test changes. Change points from the three time series are grouped in time; shading on positivity changes indicates grouping tolerance. Category labels for change point groups are shown above positivity and described in Figure 2. Black arrows indicate NPI changes; arrow direction indicates increase or decrease in stringency. For categories D and E, Y(es) and N(o) in boxes below positivity indicate whether there is a co-occurring NPI change inverse to the change in slope of positivity.

### 2.1. Data

We use national-level case and testing data as well as records on national policies for testing and NPIs (Roser et al. 2020; Hale et al. 2020). We calculate test positivity by dividing cases by tests. Testing policy is indicated by ordinal values: zero indicates no testing policy; one indicates testing of those with symptoms who meet specific criteria (e.g. known contact with a positive individual); two indicates testing of any symptomatic individuals; three indicates open public testing. For South Africa, we also use provincial-level data on COVID-19-confirmed deaths, cases, tests, and excess mortality (Mkhize 2020; National Institute for Communicable Diseases 2020; Statistics South Africa 2020; Bradshaw et al. 2020).

We selected countries for analysis based on three conditions: available case data, available testing data, and human development index (HDI) score. Of those with data, we included the countries in the lowest third of HDI score, all of which are considered low- or middle-income in 2020-2021 by the World Bank. All data used in this research are public. Further details on data and definitions found in Appendix A.1.

### 2.2. Change point detection

#### 2.2.1. Pruned Exact Linear Time (PELT) change detection

Change point detection is a set of approaches for identifying points in time where the statistical properties of a time series change (Truong et al. 2020).

We apply change point detection to epidemiological time series (cases, tests, and positivity) and national policy time series; details in Appendix A.2. Without a priori knowledge of the appropriate number of changes, the PELT algorithm must be assigned a penalty for the number of changes to identify. In the absence of an established method for this parameterization when working across time series, we developed a novel systematic approach for penalty selection which enables comparison among time series and countries; described in Appendix A.3.

#### 2.2.2. Method validation

We apply PELT to synthetic case count data generated by the stochastic agent-based COVID-19 simulator (COVASIM), in order to test PELT as a robust method for change detection in epidemiological time series (Kerr et al. 2021). The model scenario inputs include step-wise changes in contacts per person per time which represent NPI implementation, as well as a change in testing policy from symptomatic to asymptomatic testing. The model generates a simulated time series of cases and tests per thousand people, from which we calculate a positivity time series. The change point detection methods described above are applied to the seven-day mean of the time series to align with the data smoothing used with our empirical time series.

#### 2.3. Change type categorization

Change detection identifies changes that may be related to data quality, stochasticity, and testing dynamics, in addition to epidemiological changes. We classify the likely cause of changes identified by the PELT algorithm based on the co-occurrence of changes from different time series. This categorization simplifies the interpretation of epidemiological surveillance, separates signal from noise, and enables broad comparison across countries and testing dynamics.

We combine detected change points across cases, tests, and positivity time series to create change point groups. The tolerance for temporal association is set at *±* seven days to account for seven-day smoothing and weekly data reporting practices. These change groups are then categorized as shown in Figure 2, with details of the interpretation described in Appendix B. To capture all changes that may be epidemiological, we include both categories D and E as epidemiological change in our analysis. We note that these categories are heuristically defined, however they are informed both by validation using the COVASIM simulations and a qualitative understanding of epidemiological surveillance dynamics.

**Figure 2:**
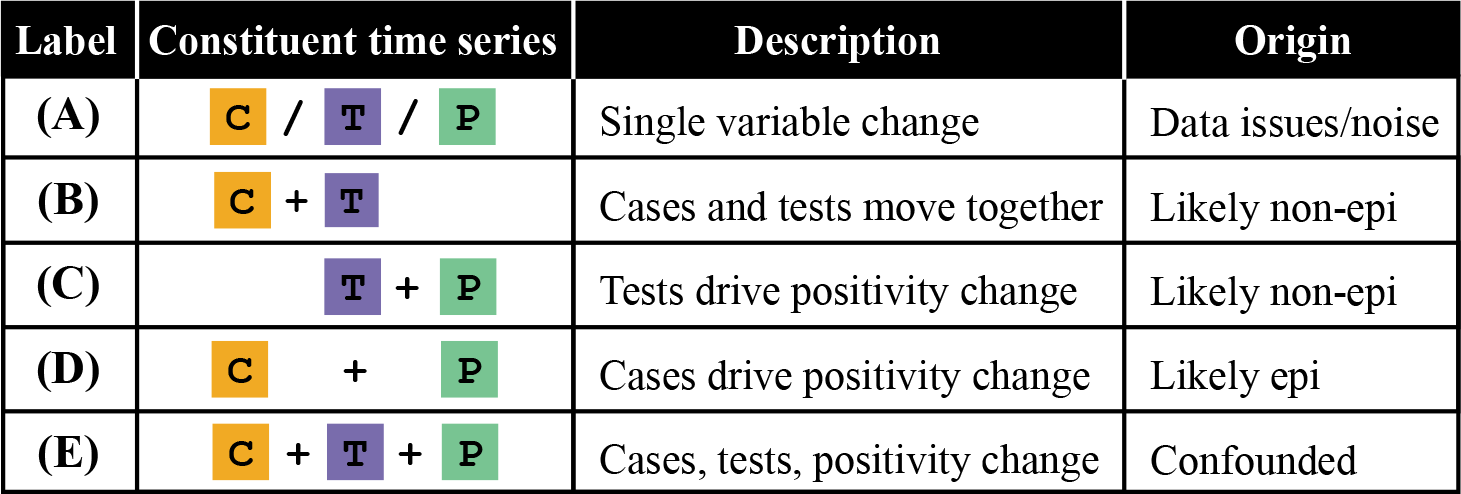
Summary of change group categories as determined by their constituent time series changes. Cases, tests, and positivity time series are indicated as orange, blue, and green, respectively. Details of the category interpretations are described in Appendix B.

### 2.4. NPI alignment

Change points classified as epidemiological are then assessed for whether they are associated with NPI changes. Timings of known NPIs in the empirical data are lagged by nine days to account for virus incubation time and the delay from symptom onset to test-seeking (Qin et al. 2020). We consider a change point to be aligned with an NPI when two conditions are met: 1) an epidemiological change co-occurs with an offset NPI and 2) the change in NPI stringency is inverse to the concurrent change in positivity slope. The second condition includes occasions when stringency was increased and positivity decreased, as well as occasions when stringency decreased and positivity increased.

## 3. Results

### 3.1. Synthetic modeling validates Pruned Exact Linear Time (PELT) is a robust method for change detection in epidemiological time series

Before applying the PELT method to the surveillance data, we validate its applicability for epidemiological systems. We apply PELT change detection to data from the transmission model described in Section 2.2.2. The sensitivity of change point detection to parameterization is illustrated in the top two case rate time series of Figure 3. The bottom positivity time series shows the detected change points for all time series, parameterized by the method described in Appendix A.3. PELT successfully identifies step changes in NPI and testing policies, as well as slope changes in cases, tests, and positivity. Further, the categories of change point groups are correctly identified in line with our classification scheme, labeled on the positivity time series in black and purple boxes and described in Figure 2.

**Figure 3:**
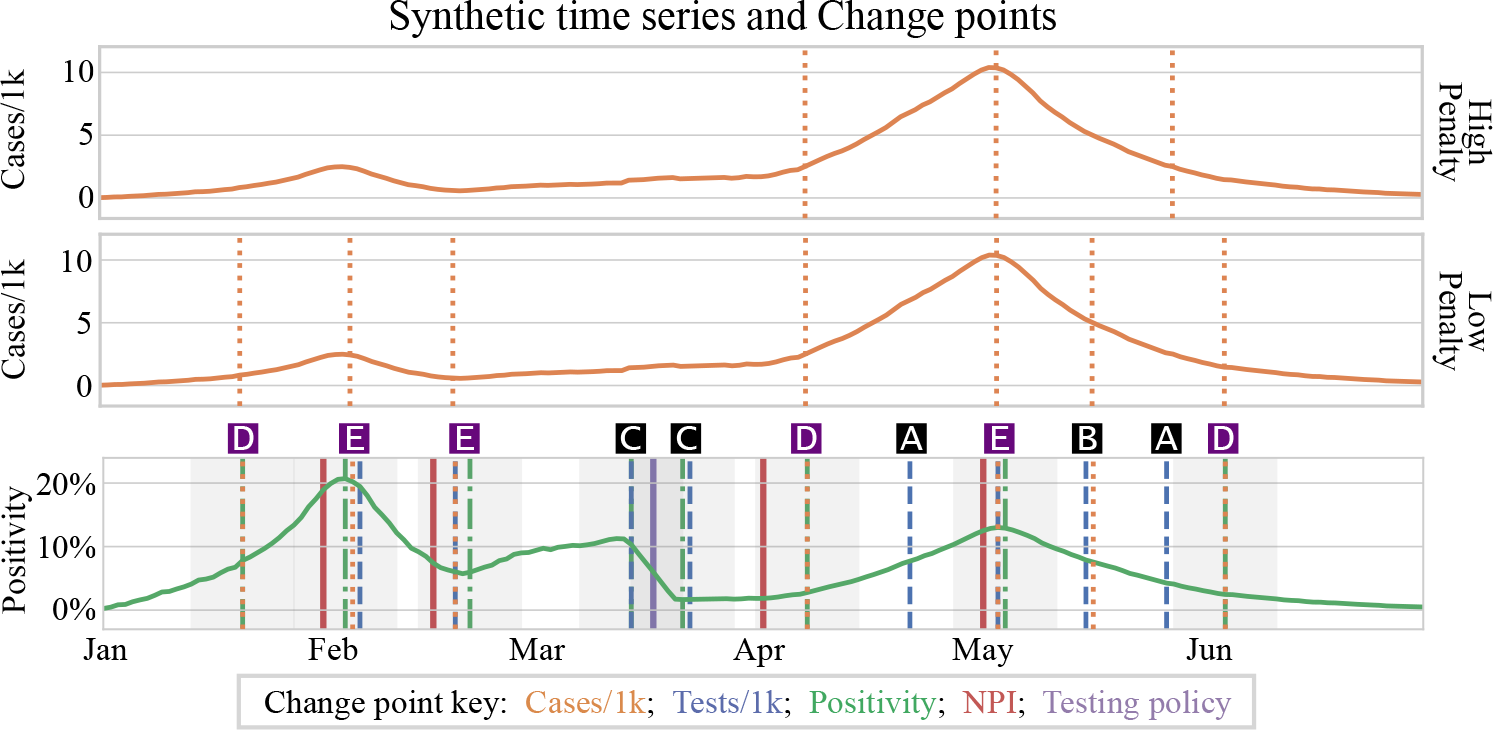
Synthetic model time series; detected change points shown as vertical lines. Upper plot shows detected change points in the cases time series using a high penalty, which promotes sparse change point detection. Middle plot shows detected cases change points in the same time series as above, but using a low penalty. Bottom plot shows positivity time series overlaid with detected change points from all time series, as indicated in the change point key. Change group categories are indicated in boxes above plot as described in Figure 2.

### 3.2. Testing rates and policies impact how surveillance measures should be interpreted

We illustrate the relevance of testing rates and the influence of testing policy using time series for Bangladesh in the context of local events (Figure 1). Cases peaked in early July, an apparent epidemiological turning point if cases were considered alone. Simultaneously, however, there was a new policy implemented to charge for testing and thus a decline in testing (Cousins 2020). This resulted in no change in positivity and contradicts the interpretation of the case reduction as a declining outbreak. Similarly, the dip in case rate in early August was accompanied by a dip in testing during the Eid al-Fitr holiday; again there is no change in positivity.

While this recommends positivity as a surveillance metric instead of case counts alone, further consideration of testing policy complicates the picture. Test eligibility in Bangladesh is based on symptoms rather than open testing, meaning that positivity is influenced by the prevalence of both COVID-19 and other respiratory illnesses. This limits the potential for positivity to detect epidemiological changes, and indeed, the positivity curve for Bangladesh is largely flat. An elaboration of COVID-19 surveillance considerations appears in Appendix C.

### 3.3. Epidemiological change detection is more influenced by testing policy than by testing rate

For all 31 LMICs in our dataset, we apply PELT change detection and change point categorization. We quantify surveillance system efficiency as the percentage of all detected change points classified as epidemiological, i.e. epidemiological change detection rate. We compare linear fits of epidemiological change detection by tests per thousand people and by testing policy (Figure 4). Results indicate that the ability to identify epidemiological changes has a stronger relationship with testing policy than with tests per thousand people. Open testing is the only testing policy bin with a mean or median epidemiological change detection rate as high as 50%, but with a wide range, indicating that open testing policy is necessary but not sufficient for quality surveillance (with outlier exceptions).

Further, LMICs have the testing capacity to measure prevalence with precision. Based on the 95th percentile of their daily testing rates, nearly all LMIC countries could measure down to 1% prevalence with a margin of error no larger than *±*1% if testing were randomly sampled (Figure 5). Only three countries hover around the margin of error to prevalence ratio of one:

**Figure 4:**
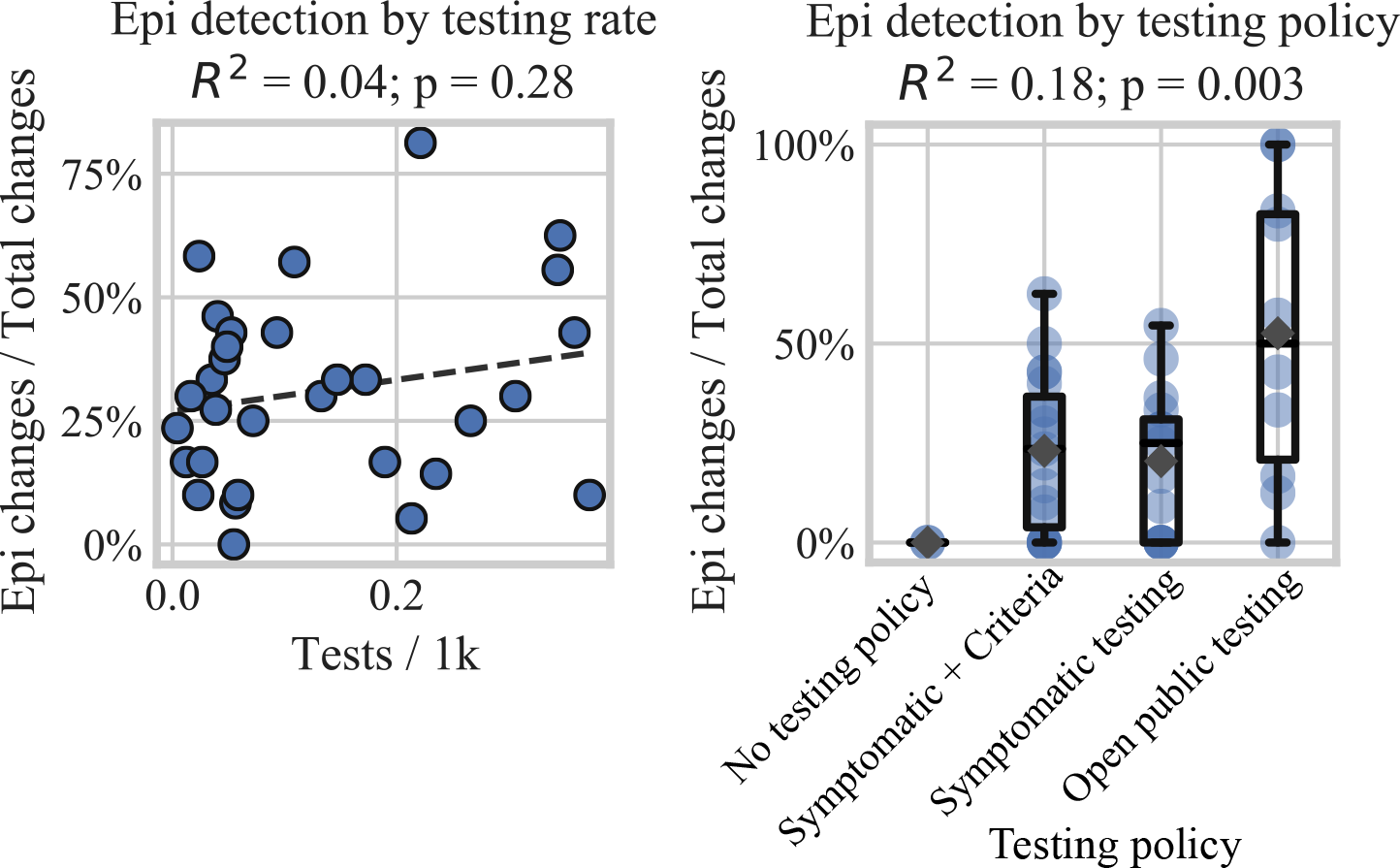
Percent of detected changes categorized as epidemiological for each country by tests per thousand people (left) and binned by testing policy (right) at the time of change detection. Linear regression shown as dotted line on left. Box and whisker plots on right show quartiles, range, and median with means plotted as gray diamonds. Note that binned calculations cause the maximum epidemiological change detection rate to differ between the two plots.

**Figure 5:**
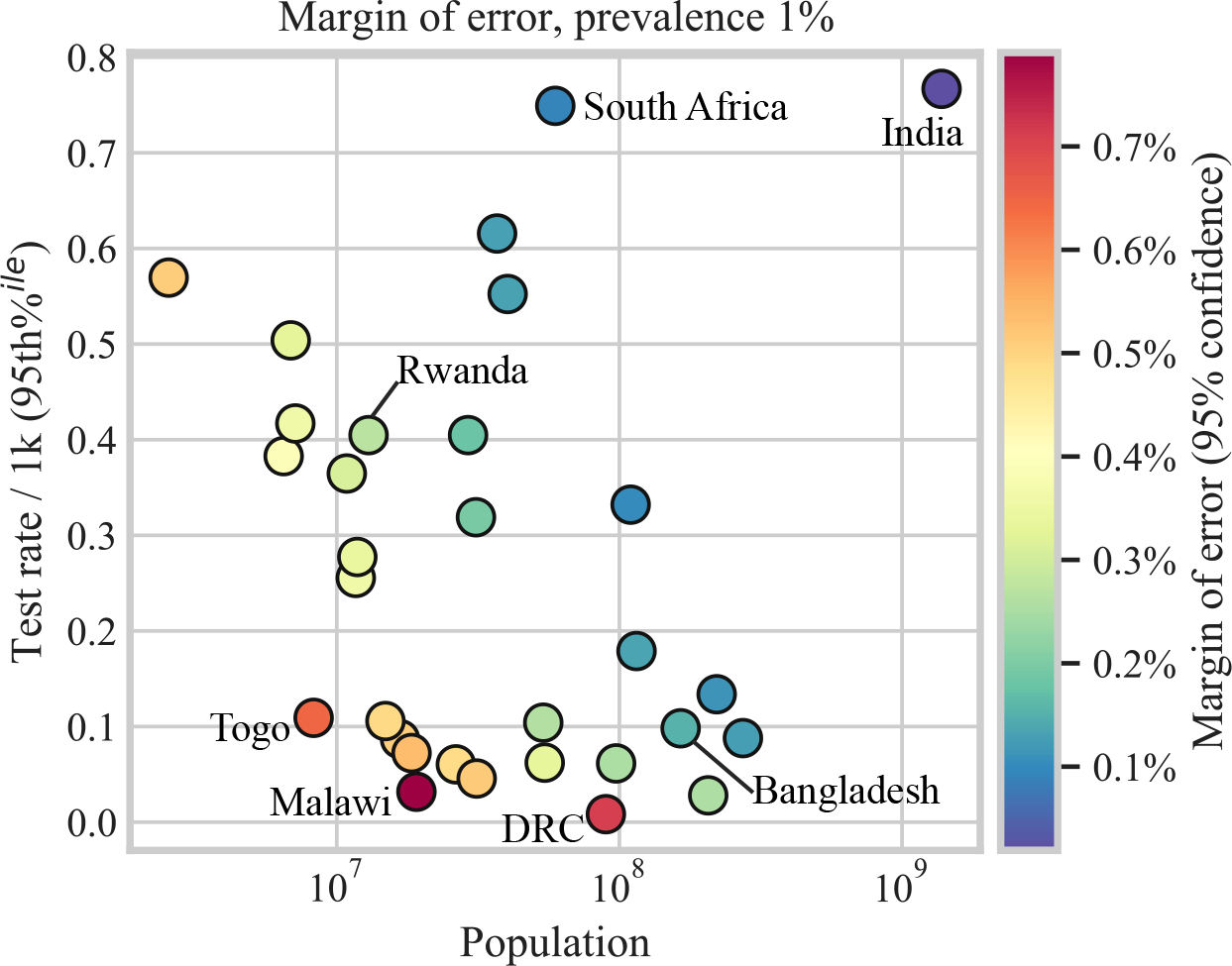
Margin of error for random sampling of 1% prevalence plotted by 95th percentile of national testing rate and population of each country in our dataset. See Appendix D for details on standard error calculations.

Malawi, the Democratic Republic of Congo, and Togo. Note that true random sampling is difficult to achieve in any setting, but open testing policies can approximate random sampling more closely than symptomatic testing.

### 3.4. Change detection rates and NPI alignment frequency vary across LMICs

Figure 6A shows the wide variation of epidemiological change detection rates across LMIC countries, with Rwanda the highest and Ethiopia the lowest. The percentage of NPIs that are aligned with a detected epidemiological change is shown in Figure 6B, again led by Rwanda. Rwanda performs well by these metrics regardless of change detection parameterization, Appendix A.4. Nearly all countries in our analysis show at least one detected epidemiological change. Conversely, approximately half of the countries in our analysis show zero alignment of any type of NPI with an epidemiological change, although the number of NPIs implemented in these countries spans a wide range.

**Figure 6:**
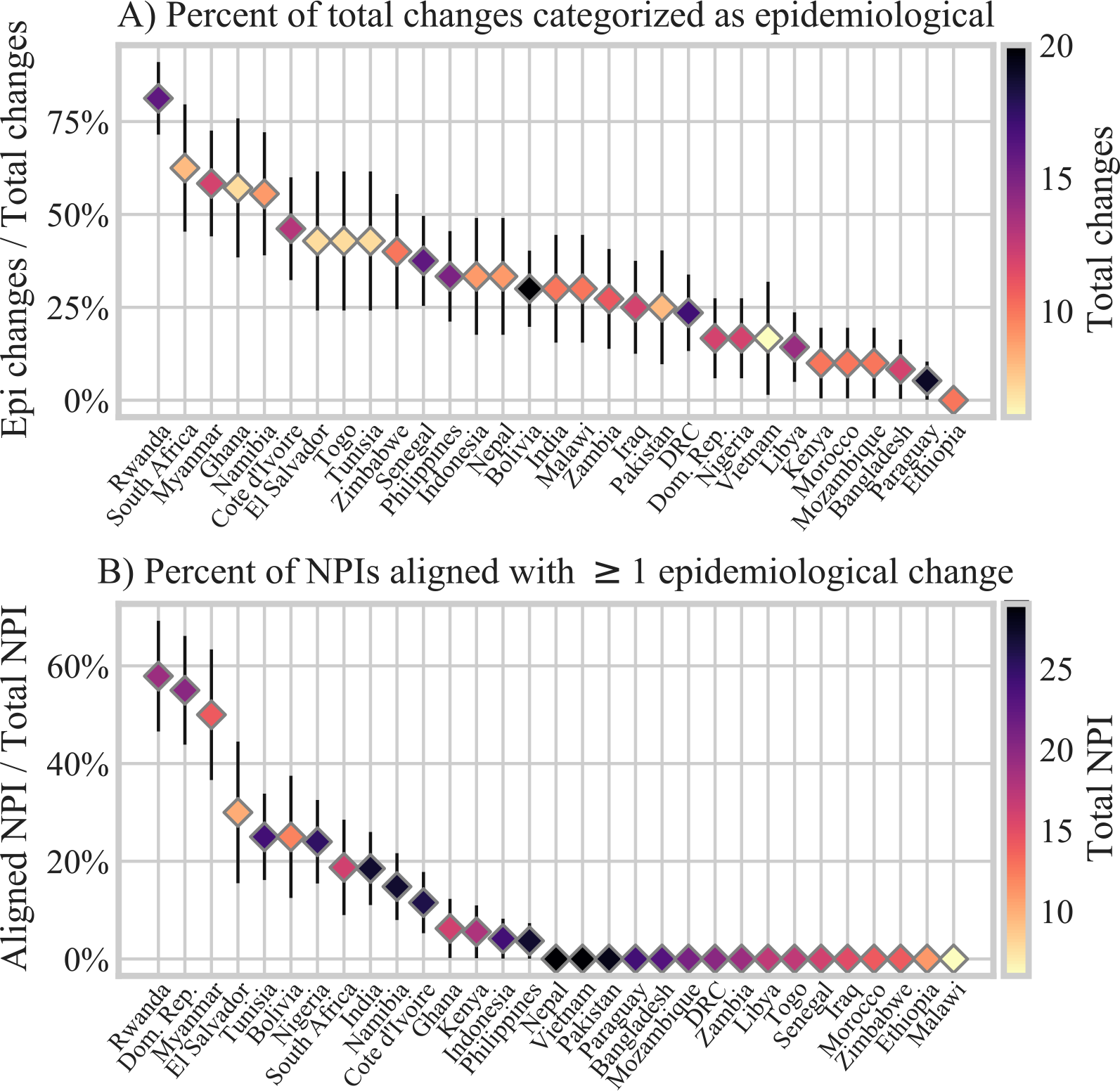
Epidemiological change detection rates (A) and NPI alignment rates (B) by country.

### 3.5. NPI alignment with detected epidemiological changes is bimodal and significant

We test the significance of NPI alignment with detected epidemiological changes through comparison with alignment rates when NPIs are assigned a random date. We find all types of NPIs measured in this study have significant rates of alignment with epidemiological changes when the zero-alignment mode is excluded (max p-value = 1.38e-10). We calculate the distributions of random NPI alignment by re-assigning random dates to NPIs by type and then finding alignment rates for N=150 bootstrapping. Across NPIs, the rate of random NPI alignment with epidemiological change has a mean of 11.6% and a standard deviation of 3.64% (gray violin distributions, Figure 7). When analyzed by country, nearly all NPI alignment rates are either higher or lower than the random date distributions (cyan circles, Figure 7). This indicates two modes of detected NPI alignment. Excluding the mode of zero NPI alignment, mean NPI alignment ranges from 50% for restrictions on internal movement, to 33% for restrictions on gathering (black squares, Figure 7). Differences in alignment rates between NPI types are not significant. Potential differences in NPI alignment rates are confounded by synchronous implementation of NPIs, although there is some evidence to support the effect strength of workplace closing and stay at home requirements (Appendix E).

**Figure 7:**
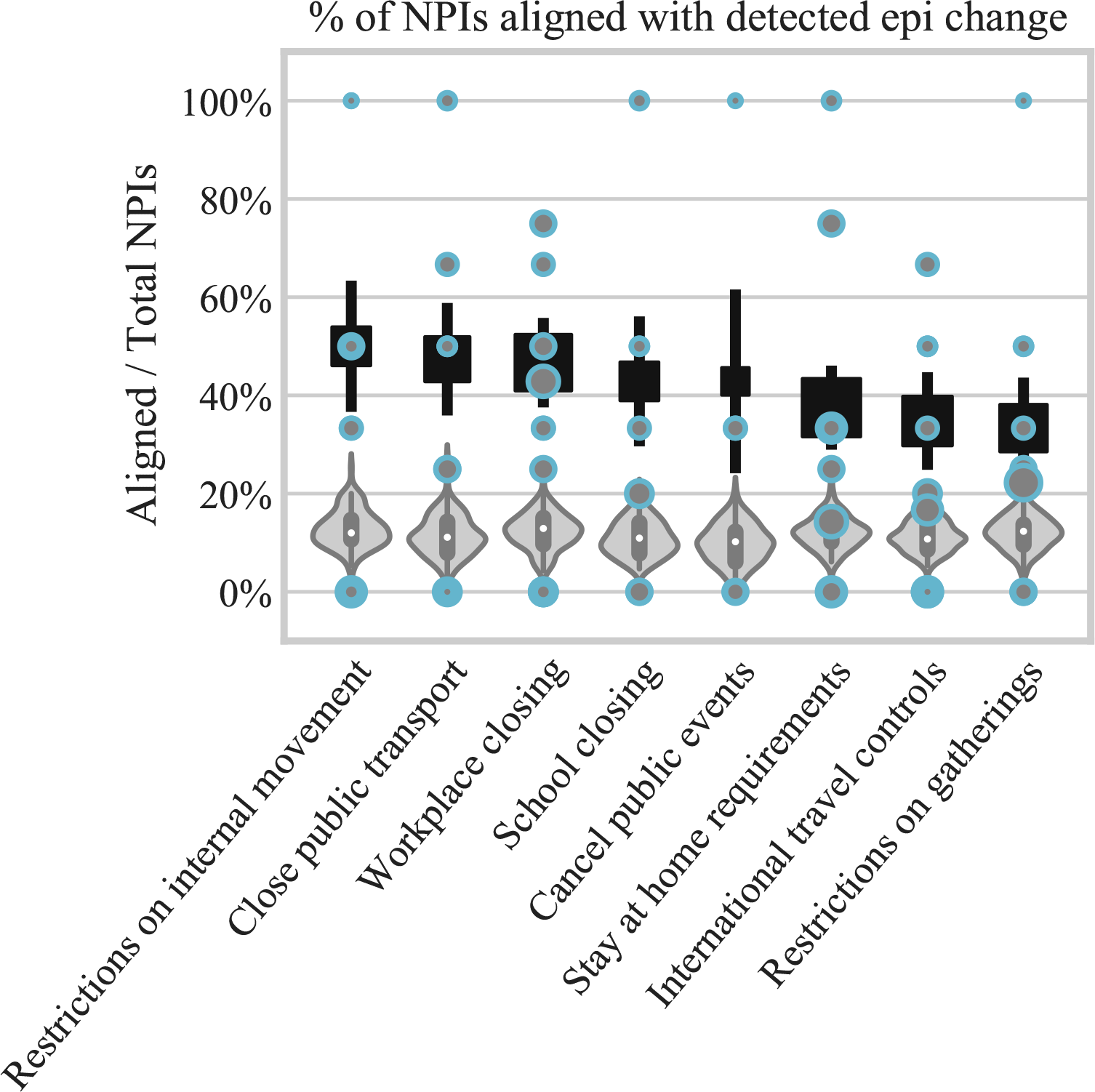
Percentage of each type of NPI that is aligned with a detected epidemiological change. Cyan circles are data for individual countries, sized by total number of NPIs by type and country. Gray violins are distributions of NPI alignment for random NPI dates. Black squares indicate weighted mean of country data for NPI alignment greater than zero; error bars indicate standard error. NPIs include both easing and tightening of policy restrictions.

### 3.6. National-level results obscure sub-national heterogeneity in epidemiological dynamics and surveillance

To investigate sub-national heterogeneity, we conduct the same analyses as above, but at the province level in South Africa. Figure 8A shows substantial variability in provinces both by NPI alignment rate and by epidemiological change detection rate. In line with results from national-level data, epidemiological change detection rate is not correlated with mean tests per thousand people. Because of reporting limitations, the NPIs here are national policies only.

**Figure 8:**
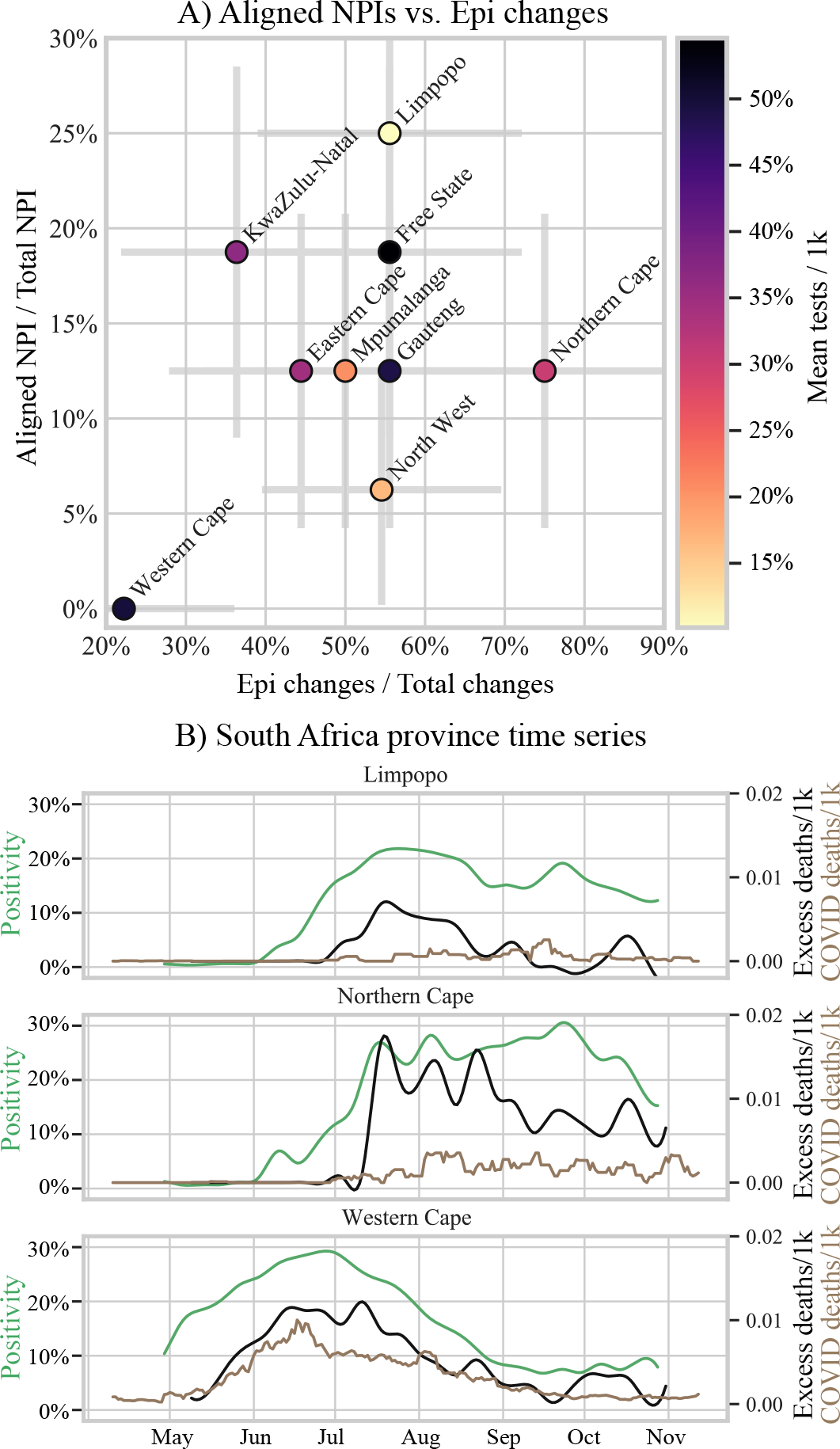
A) South African provinces by aligned NPI fraction versus epidemiological detection rate with color indicating the mean over time of tests per thousand people. B) Time series from three example provinces. Positivity shown in green on left y-axis. Deaths per thousand people shown on the right y-axis: excess mortality in black; COVID-19-confirmed deaths in brown.

We select three edge cases from the scatter plot in Figure 8A (Limpopo, Northern Cape, and Western Cape) to compare time series of positivity, COVID-19-confirmed deaths, and total estimated excess mortality (Figure 8B). The differences in the timing and trajectories of the time series illustrate strong sub-national variability in underlying epidemiological dynamics that are may be overlooked when time series are aggregated to the national level.

Variation among provinces in the difference in magnitude between excess mortality and COVID-19 deaths points to differences in their surveillance systems. Western Cape is the only province where the magnitude of excess deaths resembles that of COVID-19-confirmed deaths throughout the time series. In Northern Cape, the peak of excess deaths is roughly a factor of three higher than the COVID-19-confirmed deaths, suggesting substantial under-reporting.

## 4. Discussion

We have demonstrated a standardized and quantitative approach to analyzing epidemiological surveillance time series that can be automated for improved interpretation and comparison between countries. We find that interpretation of epidemiological trajectories are more informative when cases are normalized by tests and highlight the disadvantages of symptomatic testing for outbreak tracking and public health purposes. These findings align with literature emphasizing the importance of positivity and test sampling strategies (Pearce et al. 2020; Hilborne et al. 2020). Our finding of strong alignments of NPIs with epidemiological changes is consistent with existing literature on global NPI impacts (Islam et al. 2020; Haug et al. 2020; Liu et al. 2020). When we apply our analysis of change types to evaluate the efficiency of national surveillance systems, we find that Rwanda stands out as a country with a strong surveillance system, which is consistent with current qualitative evaluation (WHO Regional Office, Africa 2020).

Our approach substantially broadens the scope of previous analyses of COVID-19 surveillance data in LMICs. We use statistical change detection methods on COVID-19 surveillance time series from 31 LMICs to differentiate epidemiological changes from changes related to stochasticity, data quality and non-epidemiological dynamics. This maximizes the insights gained from limited data, reduces erroneous interpretations of epidemiological time series, and enables quantitative comparisons of disease surveillance approaches. We use epidemiological change detection rate as a proxy for surveillance system efficiency, and show that epidemiological change detection is not as strongly associated with testing rate as with open testing policies. There is substantial variation in epidemiological and surveillance dynamics across countries and in our sub-national analysis.

There are limitations in our analysis related to the data themselves as well as our methods. Simultaneously, these data challenges are precisely the motivation for developing our methods: maximizing information with limited data. Our data are potentially biased by unmeasured factors such as fluctuations in testing capacity and undocumented population sampling strategies over time, delays and temporal uncertainty due to reporting systems, and incentives for case-finding. Defining co-occurrence when working with imprecise time series is a challenge, which we partially mitigate by considering uncertainty bounds when defining change groups. We emphasize, of course, that co-occurrence does not establish causality. In PELT change detection, the changes detected are influenced by the choice of the sparsity parameter. In a sensitivity analysis of our novel parameterization approach, however, we find that Rwanda remains the leader in surveillance system performance, regardless of the parameterization choice.

Results from this analysis highlight that surveillance data must be used carefully to ensure proper programmatic responses. As a sufficient and less resource-intensive approximation of random sampling, open testing would enable better estimation of disease prevalence and examination of NPI impacts in geographies without reliable hospitalization data, death records, or seroprevalence surveys. NPIs without epidemiological changes may indicate inefficacy of policy, but may also indicate shortfalls of surveillance systems, which undermines policy makers’ ability to make evidence-based decisions. Our methods could be further developed and applied not just to COVID-19 but also to surveillance interpretation for other poorly measured diseases, enabling more informed decision-making and targeted improvements in surveillance systems.

## 5. Declarations

### 5.3. Ethical approval

Not required.

## Data Availability

All data used in this research are public.

## Appendix A. Methods

### Appendix A.1. Data and definitions

The case rate is defined as the number of individuals confirmed positive for the SARS-CoV-2 virus per population, regardless of symptoms. The testing rate per population is defined as the number of people tested (i.e. excluding duplicate confirmatory tests) divided by the population, regardless of the test outcome. To address the dependence of case rate on testing rate, we normalize case counts by the number of tests conducted, creating the alternate metric of test positivity rate.

For the purposes of comparing between countries and over time, we define the ‘mean testing policy’ as the average over time of the ordinal value representing the national testing policy. Thus, lower values represent more restricted testing over longer periods of time. Social distancing policies tracked in the dataset include the following: closing schools, closing workplaces, cancelling public events, restricting gathering sizes, closing public transport, stay at home requirements, restricting in-country mobility, and restricting international travel.

Weekly cases, testing, and death data are interpolated using a cubic spline. All daily cases, testing and death data are smoothed using a centered seven-day rolling average. Error bars on plots show standard error.

### Appendix A.2. PELT change detection

The naive approach to generating an exact solution to time series segmentation is to test all possible solutions. For an unknown number of changes, this also requires testing a sufficiently large set of possible number of changes. We use the Pruned Exact Linear Time (PELT) change detection method to address these computational tractability issues.

PELT minimizes the sum of costs from a criterion function across time series segments while balancing model complexity by implementing a linear penalty function and change point pruning. At each iteration of cost minimization for a potential set of change points, time points that cannot be a global minima are removed from future consideration. The PELT method, developed with applications in genetics and finance in mind, is increasingly used for climate and epidemiological applications (Killick et al. 2012; Sissoko et al. 2017; Ouedraogo et al. 2018).

To detect changes in slope of the epidemiological time series, we use the first derivative as input for the PELT algorithm. For detection of discrete step changes of policy time series, we feed the data directly into the change detection algorithm without taking a derivative. For all time series, we use the radial basis function kernel for the PELT detection algorithm.

### Appendix A.3. PELT parameterization

To date, there is no established method for parameterizing the PELT change density penalty across time series when the number of changes is not known. One of the ways to choose penalty values across time series would be to unify the number of changes detected in each time series. This, however, imposes the assumption that all time series exhibit the same general change frequency and it is only the point in time of a change that is unknown, rather than the number of changes.

We present here a novel approach for systematic parameterization when identifying an unknown number of changes in slope over many time series, as in our case with multiple epidemiological time series across countries. To accomplish this, we first conduct change detection in a sweep over parameter space. The change points detected using a given value in parameter space slice the time series into segments, each of which is input into a linear regression. The standard error for each of those linear regressions is calculated and then averaged, weighted by segment length.

The mean standard error associated with each penalty value, when plotted over parameter space, is characterized by a series of plateaus that correspond to plateaus in the number of changes found with each penalty value, Figure A.9, top row. Descending through penalty values in the penalty parameter space, the lowest penalty associated with each plateau is selected to represent that plateau.

Each time series is thus associated with a sparse set of penalty values, ordered from largest penalty (low change point density) to smallest penalty (high change point density). The penalty values are unique to each time series, but represent the same ordered progression of plateaus. To illustrate, change detection with different ranked penalties for South Africa and Bangladesh are shown in green in Figure A.9.

Penalty values for each unique time series can then be chosen based on their order in the ranked plateau list. This enables a principled approach to parameterization that creates change density parity across time series, allowing for the likelihood that some time series are characterized by more changes than others. Among all time series and countries in our analysis, the minimum number of plateaus detected is four. We therefore choose the fourth penalty value for all time series.

**Figure A.9:**
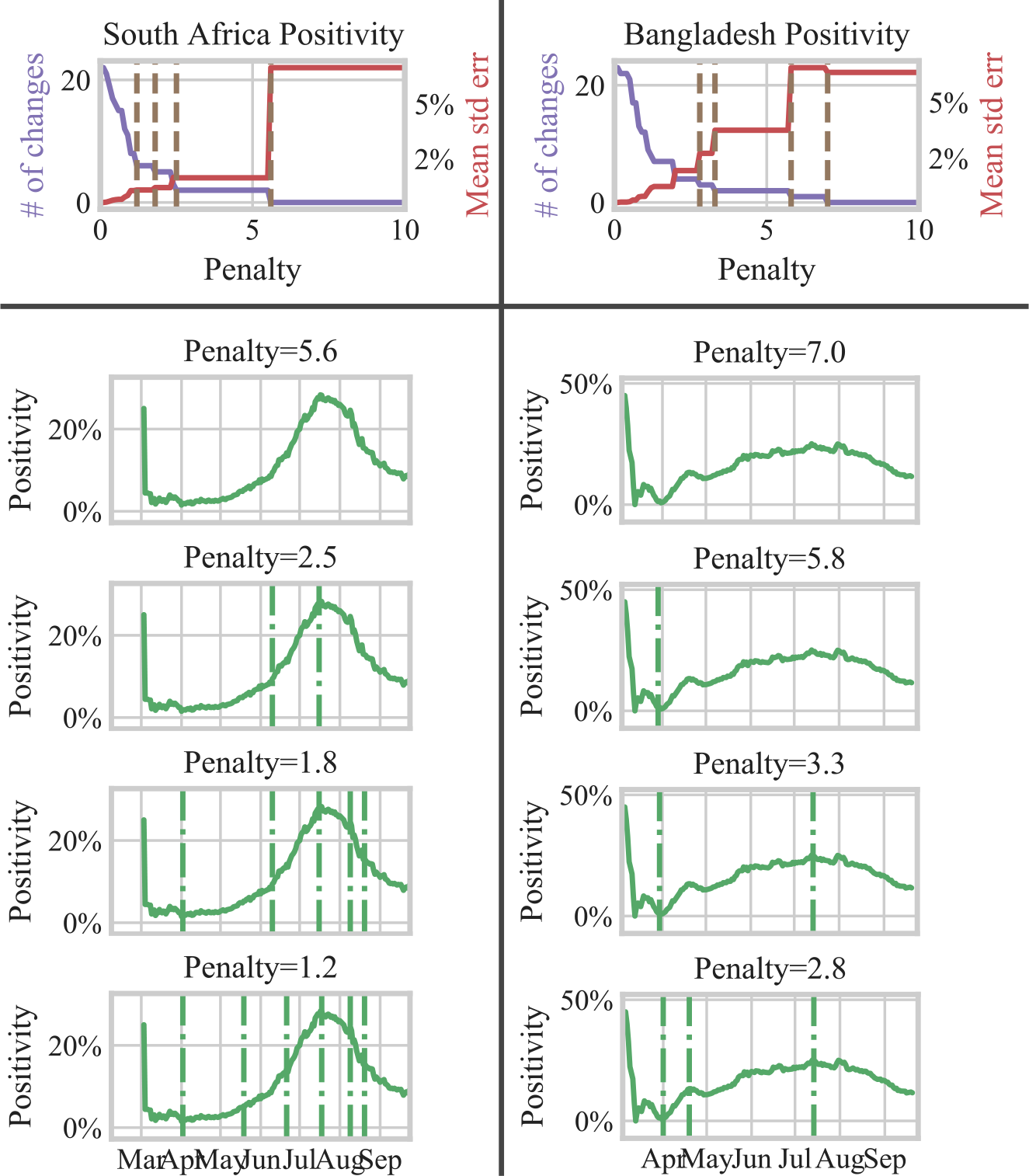
Top row: change detection results over parameter space for the positivity time series of South Africa (left) and Bangladesh (right). For each penalty value, the associated number of changes is plotted in purple on the left y-axis and the mean standard error of linear regressions of the time series segments are plotted on the right y-axis. The parameter values selected to represent plateaus are shown as brown dotted vertical lines. Bottom row: positivity time series for South Africa (left) and Bangladesh (right) plotted with detected changes as vertical lines for each of the four penalty values selected to represent plateaus in the top row.

#### Appendix A.4. Parameterization sensitivity analysis

To evaluate the influence of penalty selection on our analysis results, we conduct a parameterization sensitivity analysis. We compare results of country ranking by epidemiological change detection rate for different penalty plateau selections. Skipping penalty rank one for which no changes may be detected (see examples in Figure A.9), we find that regardless of which penalty rank we use, Rwanda appears at the top of the list with the highest epidemiological change detection rate.

**Figure A.10:**
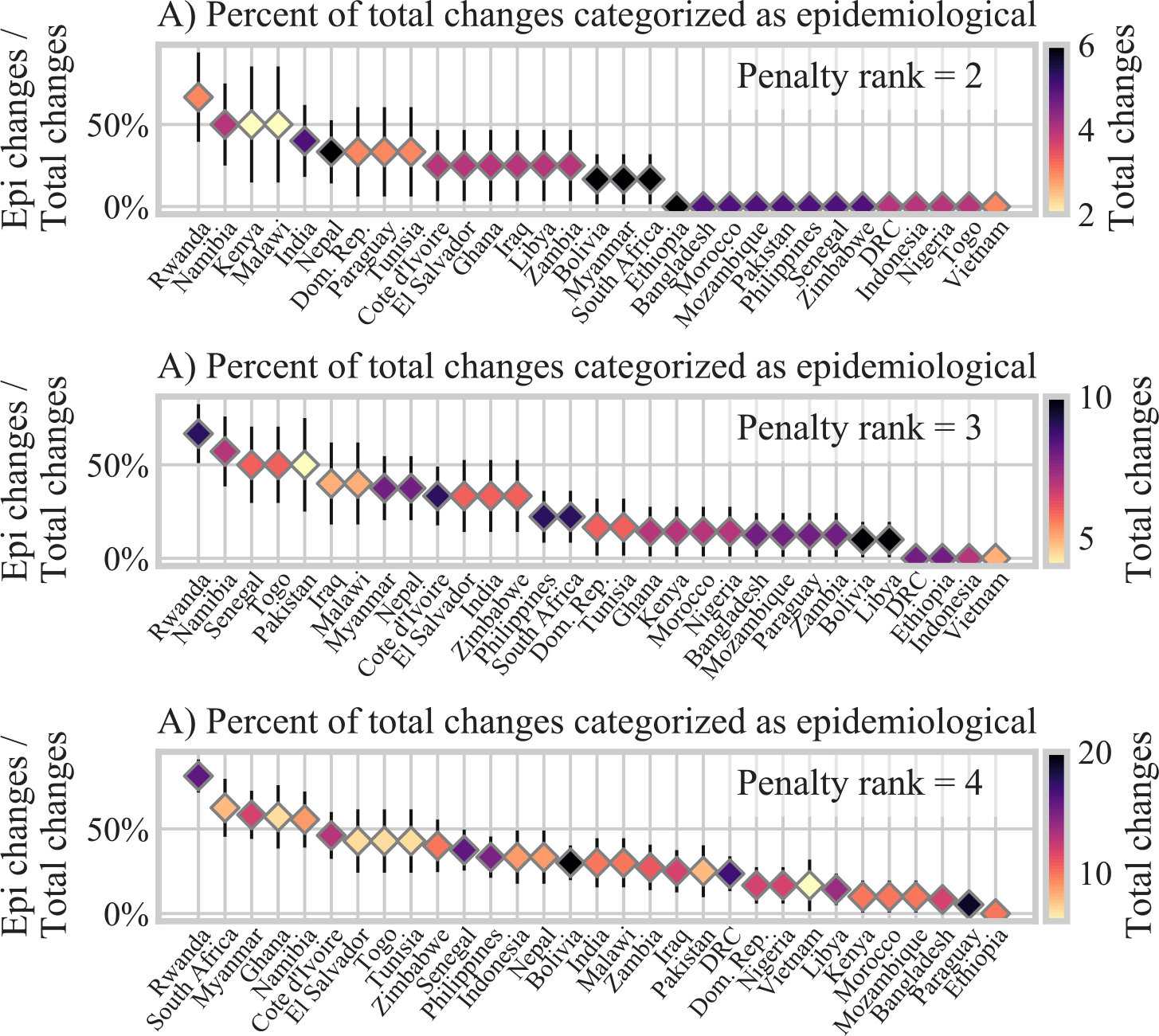
Countries ranked by epidemiological change detection rate, as in Figure 6, shown for different choices of penalty parameterization. Order top to bottom follows order of time series of Figure A.9, without rank one for which there are often no changes.

## Appendix B. Change categorization

### Appendix B.1. Heuristic interpretation

Detected change points across cases, testing, and positivity time series are combined into groups by temporal co-occurrence. These groups are then categorized by their constituent time series, Figure 2. Dynamical interpretation of the constituent time series aids in the characterization of each change group category, as follows:

A) Single variable change: Because positivity is defined according to the arithmetic relationship, *Positivity* = *Cases/Tests*, a change in any one of the variables should be accompanied by a change in at least one of the other variables. A single change in only one of the variables indicates that the change arises from issues in the data or noise. These single variable changes often occur early in the time series, when the numbers of cases and tests are smaller, signal to noise ratios are lower, and confidence intervals are larger.
B) Cases and tests change: Tests and cases move up or down together. What might look like a significant change in cases is associated with a change in testing, likely not a change in epidemiology. Factors affecting testing include testing capacity, care-seeking behavior, and testing sampling policy. With this change category, the change in testing could be a change in capacity or care-seeking, but the lack of change in positivity indicates testing is still sampling the same population the same way, without changes in epidemiology.
C) Tests and positivity change: Positivity change is driven by testing change, not a change in cases. An increase or decrease in testing does not impact absolute numbers of detected cases, which suggests a change in test sampling. Dynamics that would produce this pattern include, for example, adding population with lower prevalence in the case of open testing, or limiting testing to a higher-prevalence population in the case of symptomatic testing. It is also possible, however, that a change in testing sampling masks a simultaneous change in epidemiology. In this situation, the change in testing would have to precisely offset the change in epidemiology to observe this category of change association. Category C is thus designated to likely indicate a non-epidemiological change.
D) Cases and positivity change: Positivity change is driven by a change in cases without a change in testing. This suggests a change in epidemiology, but the significance may be different under random vs. symptomatic testing. Under random testing, this type of change arises only with a change in SARS-CoV-2 epidemiology. Under symptomatic testing, the restriction of sampling to CLI means that a change in the epidemiology may be confounded by a change in CLI epidemiology. Note also that symptomatic testing captures changes only in symptomatic SARS-CoV-2 (i.e. cases of COVID-19). Another possible explanation for this combination of changes is a change in sampling without a change in the absolute number of tests. This might occur, for example in a switch from symptomatic to open testing. For this reason, we categorize this change combination as likely instead of certainly epidemiological.
E) All three variables change: With a change in cases, tests, and positivity, it remains difficult to disentangle epidemiological from non-epidemiological factors. Category E can be considered a combination of categories C and D, and the testing and case changes may or may not be independent. To capture all changes that may be epidemiological, we consider categories D and E to be epidemiological changes, and categories A, B, and C to be non-epidemiological changes.

A principal component analysis (PCA) supporting the separability of change categories is detailed in Appendix B.2.

### Appendix B.2. PCA analysis of change categories

**Figure B.11:**
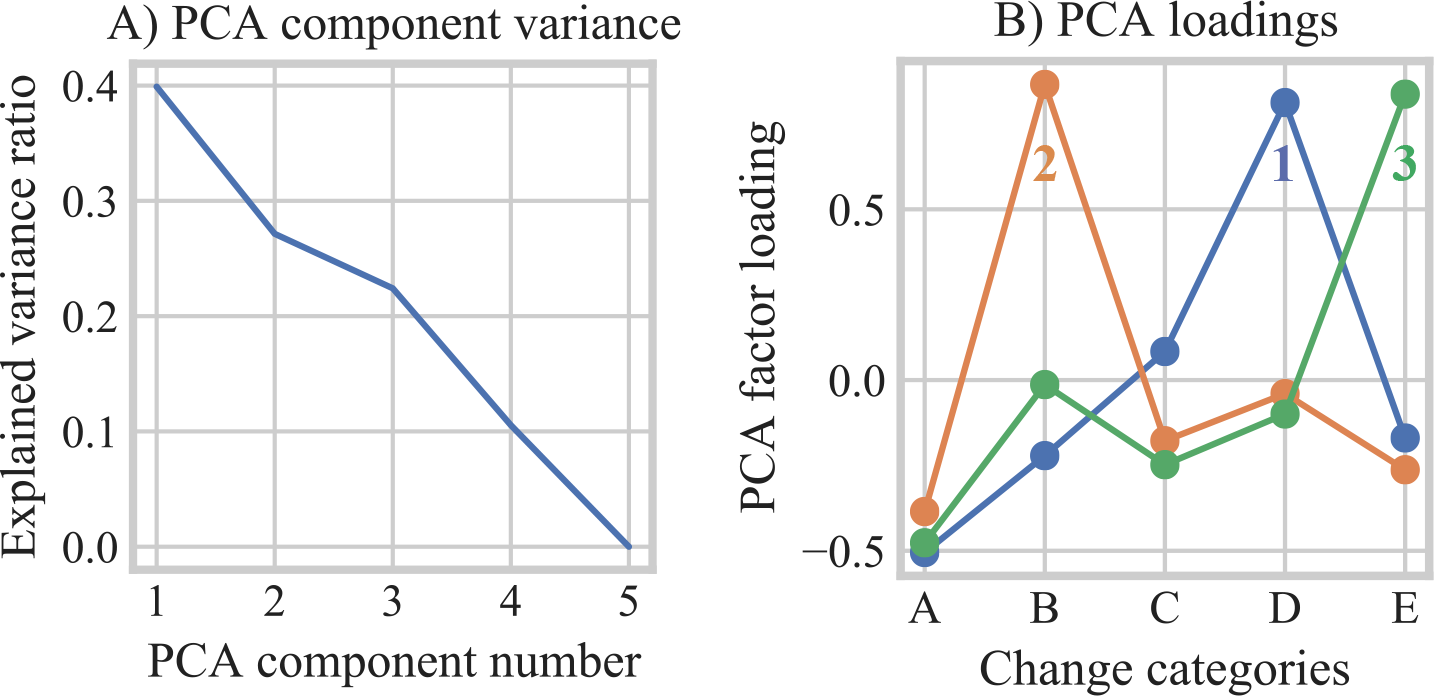
PCA of countries by change category detection rate (i.e. number of changes in each category divided by total changes detected for each country). A) Explained variance ratio by PCA component number; B) PCA factor loadings by change categories.

In addition to the dynamical interpretation of constituent time series (Appendix B), we show the separability of change categories with a principal component analysis (PCA). The surveillance results of different countries are quantitatively characterized by a PCA of the relative frequency with which they detect different categories of changes. The PCA establishes how categories do or don’t represent axes of difference across countries.

Based on the curve of explained variance ratio by PCA components (Figure B.11A), we choose the first three PCA components to examine factor loadings (Figure B.11B). Each component is dominated by a single category, in PCA component order: category D (epidemiological change); category B (testing artifacts); and category E (confounded). Each of these PCA components is anti-correlated with category A (noise). These relationships among the different change categories is consistent with our dynamical interpretation. Figure B.12 shows the frequencies of change categories for those categories that dominate the factor loadings for all countries in our dataset.

**Figure B.12:**
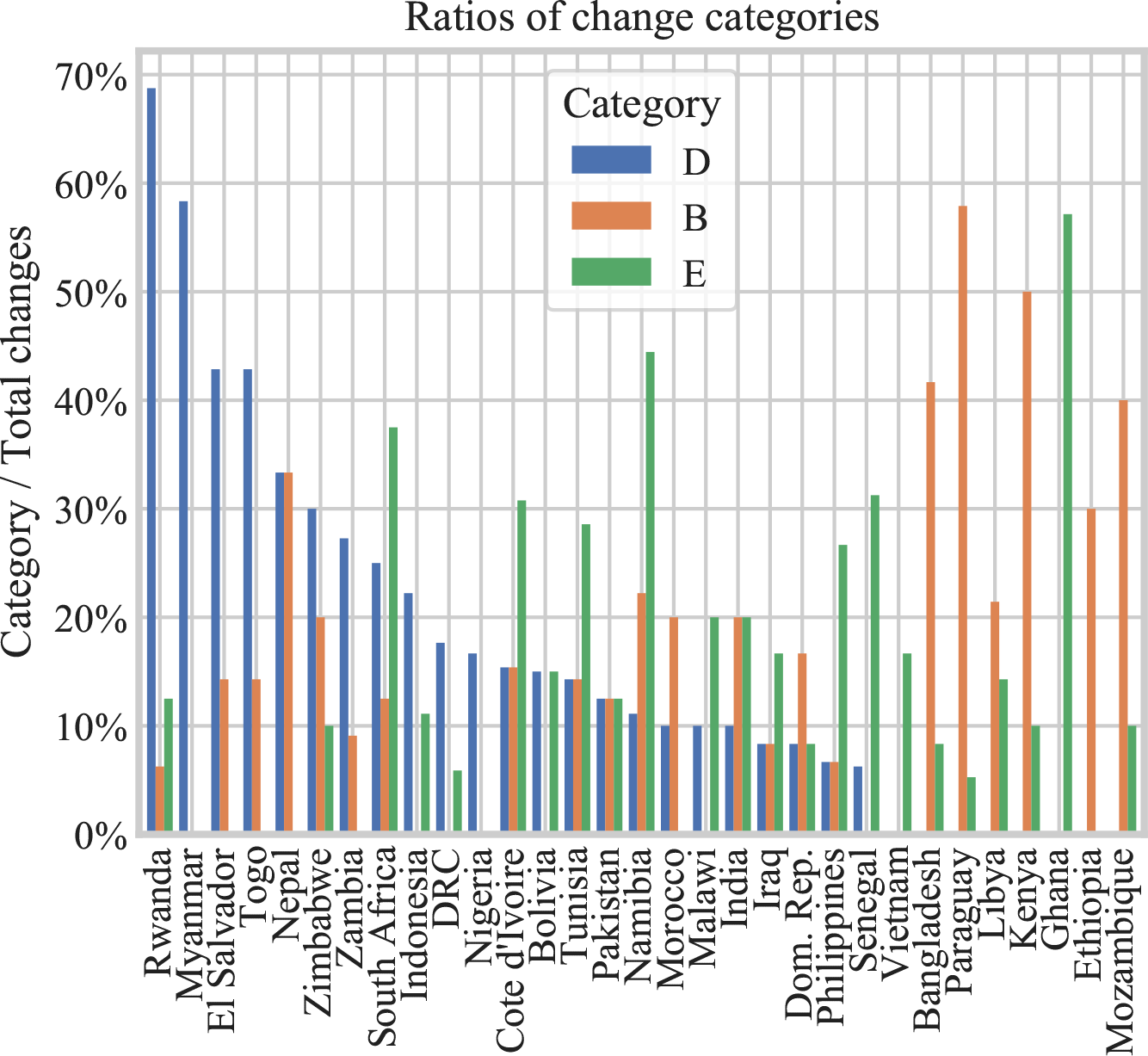
Change category frequencies by country for categories D, B, and E, chosen according to the dominant categories in the PCA factor loadings.

## Appendix C. Surveillance considerations

Below we lay out in basic terms considerations for three components of SARS-CoV-2 epidemiological surveillance: population, testing, and their role in surveillance metrics. We demonstrate that the testing strategy of random testing with the surveillance metric of positivity is the combination that best represents SARS-CoV-2 prevalence. Here we use the terminology of SARS-CoV-2 to include all asymptomatic and symptomatic infections.

Population is composed of people with and without SARS-CoV-2. Of those with SARS-CoV-2, some are asymptomatic, some are symptomatic. Of those without SARS-CoV-2, some are non-symptomatic, others have symptoms of non-COVID-like-illness, and some have COVID-like-illness (CLI) symptoms.

Relevant components of testing include eligibility for testing under a given testing framework, as well as testing rate and capacity. Under random sampling, the general population is eligible for testing; symptomatic testing restricts eligibility to people with CLI symptoms. Testing rate is a measure of tests conducted per total population, while testing capacity indicates the proportion of eligible individuals who are actually tested.

Detected cases as a surveillance metric is a function of number of tests, the eligible testing pool, and the total cases within the testing pool. Positivity is defined as detected cases per tests conducted.

Applying these formulations to surveillance metrics, we can see that detected cases under symptomatic testing is not only a function of number of tests conducted, but also of the number of individuals exhibiting CLI symptoms. CLI in turn is a function of non-SARS-CoV-2 CLI and symptomatic SARS-CoV-2.

Positivity under symptomatic testing is normalized for number of tests conducted, measuring not general prevalence in the population, but the portion of CLI that is symptomatic COVID-19. Metrics derived from symptomatic testing do not account for asymptomatic SARS-CoV-2 and are confounded by non-SARS-CoV-2 CLI.

As with symptomatic testing, detected cases under random testing are a function of number of tests. The sampling, however, is taken from the general population, and thus positivity under random testing is a metric that represents prevalence.

As tests approach eligible under symptomatic testing, cases detected equals CLI COVID-19 cases. Note, however, that testing coverage (i.e. *tests/eligible*) is not only influenced by the number of tests processed, but also reporting rate. Who shows up for testing is a subset of the people who would be eligible for testing.

Assuming capacity to test all eligible individuals and perfect reporting rates, symptomatic testing would still yield only the number of symptomatic COVID-19 cases. For random testing, testing rate is equivalent to testing coverage, and case counts depends on testing. The random testing positivity metric does not depend on testing rate, and captures both symptomatic and asymptomatic COVID. The relationship shown empirically in Section 3.3 wherein increasingly open testing policies are associated with increasingly effective epidemiological change monitoring supports the equation-based result that random testing is more suited to epidemiological surveillance.

Population components:

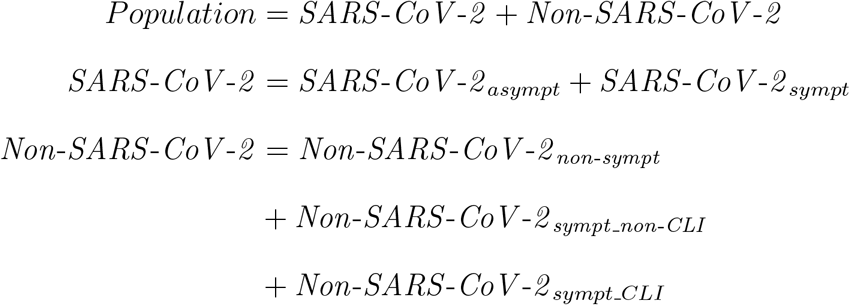

Testing components:

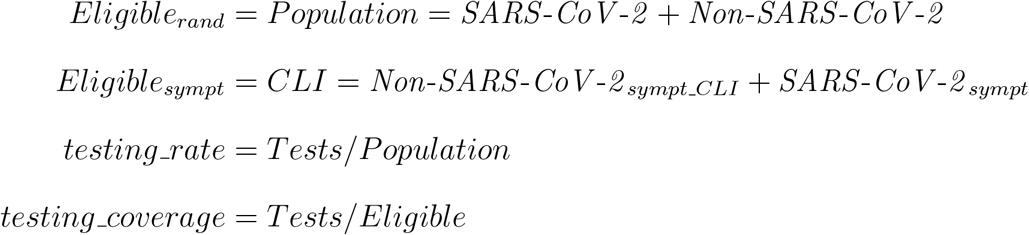

Surveillance metrics:

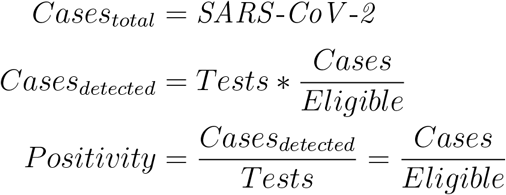

Symptomatic testing:

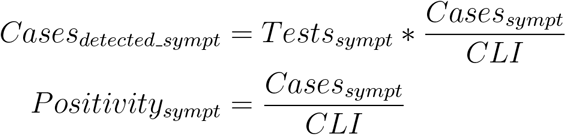

Random testing:

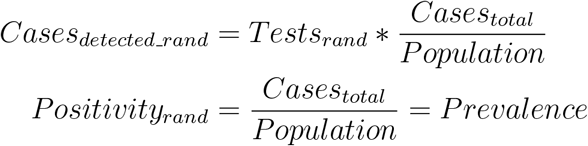

## Appendix D. Summary statistics

For the purposes of understanding the sensitivity of a given level of testing, we define the standard error for positivity, as number of positive tests per total number of tests, we calculate standard error as follows:

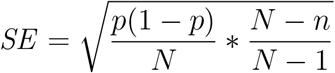

Where *n* equals total number of tests, *N* equals population, and *p* equals the number of positive tests per the total number of tests. The corresponding margin of error equals one-half the confidence interval, and when calculated at the 95% confidence level is as follows:

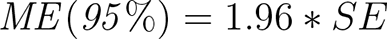

Note that this formulation of confidence interval is not reliable when number of tests is very small, or probabilities are very close to zero or one. Under the condition of true random testing, positivity is a direct measure of prevalence. At any given prevalence, margin of error can be calculated for the number of tests administered and the total population. This calculation is carried out for all LMIC countries in our dataset. Margin of error is then normalized by the given prevalence rate. Based on these relationships, *ME* (*95* %)*/Prevalence* is higher at lower prevalence. In other words, precise measurement becomes increasingly more difficult as prevalence decreases.

## Appendix E. NPI correlation

Figure E.13 illustrates aligned NPI co-occurrence. Correlation score is an indicator of how often a given type of aligned NPI is implemented simultaneously with another aligned NPI. Although alignment of an NPI with an epidemiological change can be established, in the case of co-occurrence of two or more aligned NPIs, it is challenging to separate possible effects between the two types. Nonetheless, low correlation score accompanied by high frequency may be indicative of NPIs more likely to be the dominant forcing. This is the case with stay at home requirements and workplace closing, bottom plot of Figure E.13. Conversely, high correlation associated with low frequency indicates NPIs that do not often align with epidemiological change independently from other aligned NPI types. The NPIs of cancelling public events and restrictions on internal movement are examples of this case.

**Figure E.13:**
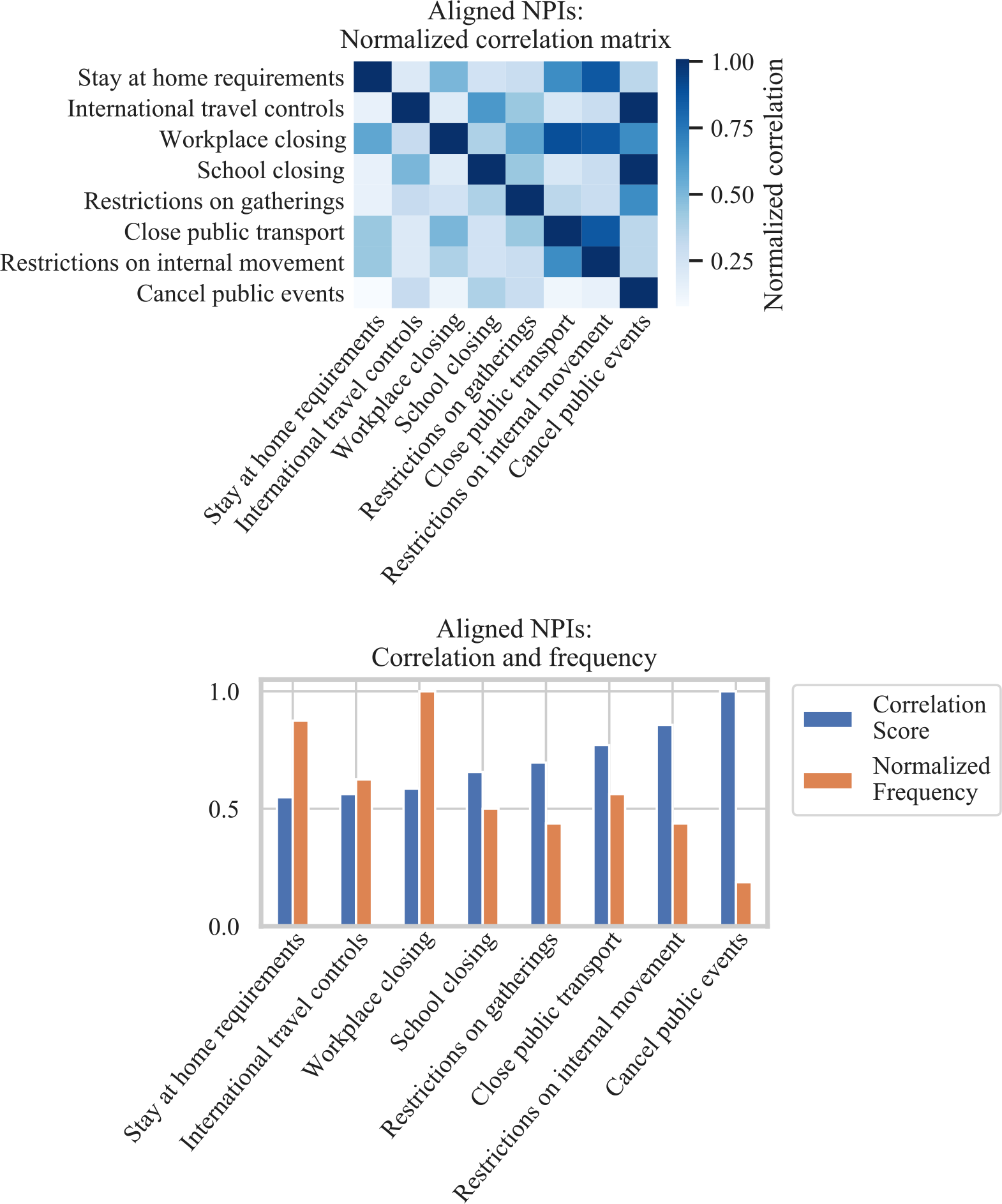
Top: Correlation matrix of co-occurrence of NPIs aligned with epidemiological changes. Correlations are normalized along the y-axis: counts of co-occurring NPIs are divided by NPI counts on the diagonal. Bottom: Correlation score and frequency of aligned NPIs by type. Correlation score is the sum of the correlation matrix along the y-axis, normalized to one. Normalized frequency is the count of aligned NPI by type normalized to one.

